# The Impact of MFN on Oncology and Hematology Treatments

**DOI:** 10.64898/2026.02.19.26346624

**Authors:** H.P. Bowen, G. O’Loughlin, C. Drake, C. Schleicher, D.G. Schulthess

## Abstract

**Background:** The Most Favored Nation (MFN) policy is a mechanism that incorporates foreign prices to determine the maximum allowable net price for any branded drug within US government-funded healthcare. Two proposed rules, the *Global Benchmark for Efficient Drug Pricing* (“GLOBE”) (90 Fed. Reg. 60,244) for Medicare Part B and the *Guarding US Medicare Against Rising Drug Costs* (“GUARD”) (90 Fed. Reg. 60,338) for Medicare Part D, invoke the Center for Medicare and Medicaid Innovation Center’s payment and service model demonstration and waiver authority, under Section 1115A of the Social Security Act (42 U.S.C. § 1315a), to calculate the US MFN price which is the lowest average price within a basket of specified foreign countries. Unlike voluntary manufacturer agreements, GLOBE and GUARD would mandate participation from all applicable manufacturers.

**Methods:** We derive MFN’s potential impact on Medicare pricing from a proprietary dataset provided by IQVIA which contained net prices for the top 37 oncology products by total US sales from January 1, 2019 through June 30, 2025 ranked by total US sales in the following countries: Australia, Belgium, France, Germany, Ireland, Italy, South Africa, Spain, Switzerland, the UK, and the US. For each drug, we select the lowest GDP-adjusted international price from a basket of those countries within 60% of the US GDP per capita, adjusted for purchasing power parity, and calculate the reduction in US price required to match its MFN price, and hence the corresponding reduction in revenues under MFN. A retrospective Net Present Value (NPV) analysis is then used to address the counterfactual question of whether each drug would have been developed had MFN pricing been in place at the time of its FDA approval.

**Results:** Under MFN, the average reduction in US prices across our drug cohort was 67%. Eighty-four percent of the 37 cancer drugs in our cohort evidenced a negative NPV if MFN had been in place at the time of their FDA approval and the commercial market is impacted. When the analysis is restricted to MFN’s impact on Medicare, the indications for these lost drugs have a total US population of 2.4 million patients. When the analysis is combined across the Medicare and commercial markets, the loss of lead indications impacts over 15 million US patients.

**Conclusions:** Mandatory MFN policies reduce the financial incentives required to develop cancer medicines; our projections show a substantial decline in new cancer drug launches and will likely lead companies to pursue indications for populations outside Medicare’s authority. If so, MFN will reduce the number of new therapies for the very population the Executive Orders are allegedly designed to aid: the Medicare-aged population who require effective new therapies in areas of high unmet medical need, such as late-stage cancers. This creates the perverse outcome of a policy nominally designed to help Medicare beneficiaries by instead redirecting innovation away from their most urgent therapeutic needs.

## Introduction

On July 31, 2025, President Trump sent letters to leading pharmaceutical manufacturers outlining the steps they must take to bring down US prescription drug prices to match the lowest price offered in other developed nations.^1^ Called Most Favored Nation (MFN), these policies were outlined in a previous Trump Administration’s Executive Order from May 12, 2025, requiring “most-favored-nation price targets to pharmaceutical manufacturers to bring prices for American patients in line with comparably developed nations.”^2^

On December 23, 2025, the Center for Medicare and Medicaid Services (CMS) published two proposed rules outlining an implementation plan for MFN titled *Global Benchmark for Efficient Drug Pricing* (“GLOBE”) (90 Fed. Reg. 60,244)^3^ for Medicare Part B, and *Guarding US Medicare Against Rising Drug Costs* (“GUARD”) (90 Fed. Reg. 60,338)^4^ for Medicare Part D. Both rules use, as justification to implement MFN pricing, the Innovation Center payment and service model demonstration and waiver authority under Section 1115A of the Social Security Act (42 U.S.C. § 1315a). In practice, CMS’s rules propose selecting drug prices from among those OECD countries whose purchasing power parity (PPP) adjusted gross domestic product (GDP) per capita is within 60% of that of the United States. A country must also have a minimum aggregate GDP of $400 billion (PPP -adjusted).^5^ Unlike voluntary manufacturer agreements, GLOBE and GUARD would mandate participation by all applicable manufacturers.

This paper examines the potential impacts of MFN pricing on the development of 37 Oncology and Hematology treatments. Cancer has had an outsized influence on U.S. biopharmaceutical R&D, as from1998 through the end of 2024, the FDA approved 217 NMEs for the treatment of oncological and hematological malignancies,138 of these drugs (64%) were approved for oncological indications.^6^ Further, only 3.4% of oncology medicines progress from Phase I to FDA approval, a stark contrast to the 13.8% average across all therapeutic areas.^7^

We construct the theoretical MFN price for each of these treatments using the lowest GDP-adjusted price in the eligible country basket. We then conduct a retrospective net present value (NPV) analysis under scenarios modeling MFN applied in Medicare alone and with spillover into the commercial market (i.e., the likelihood that commercial payers in the US demand MFN prices from pharmaceutical companies, leveraging payer coverage and formulary placement decisions).

## Data and Methods

We derive MFN’s potential impact on Medicare’s ‘best available price’ from a proprietary dataset provided by IQVIA which collects and tracks prescription cost and volume data for both gross and net pricing globally. From this data, we extracted the top 50 selling therapeutics in oncology and hematology by total US sales and approved indications from January 1, 2019 through June 30, 2025, and filtered products that were not anti-cancer interventions such as steroids, blood thinners, etc. Our remaining cohort was 37 oncologic and hematologic treatments. Drug pricing and volume data was obtained for the following countries: Australia, Belgium, France, Germany, Ireland, Italy, South Africa, Spain, Switzerland, the UK, and the US.

The lowest price — the MFN price — for each drug among the subset of countries with purchasing power parity (PPP)-adjusted GDP per capita within 60% of the US was then identified (Figure 1). For each drug, its MFN price was compared with its US price to determine the US price reduction needed to match the MFN price. These per-drug net price reductions were then applied to estimate the revenues expected under MFN pricing.

**Figure 1:**
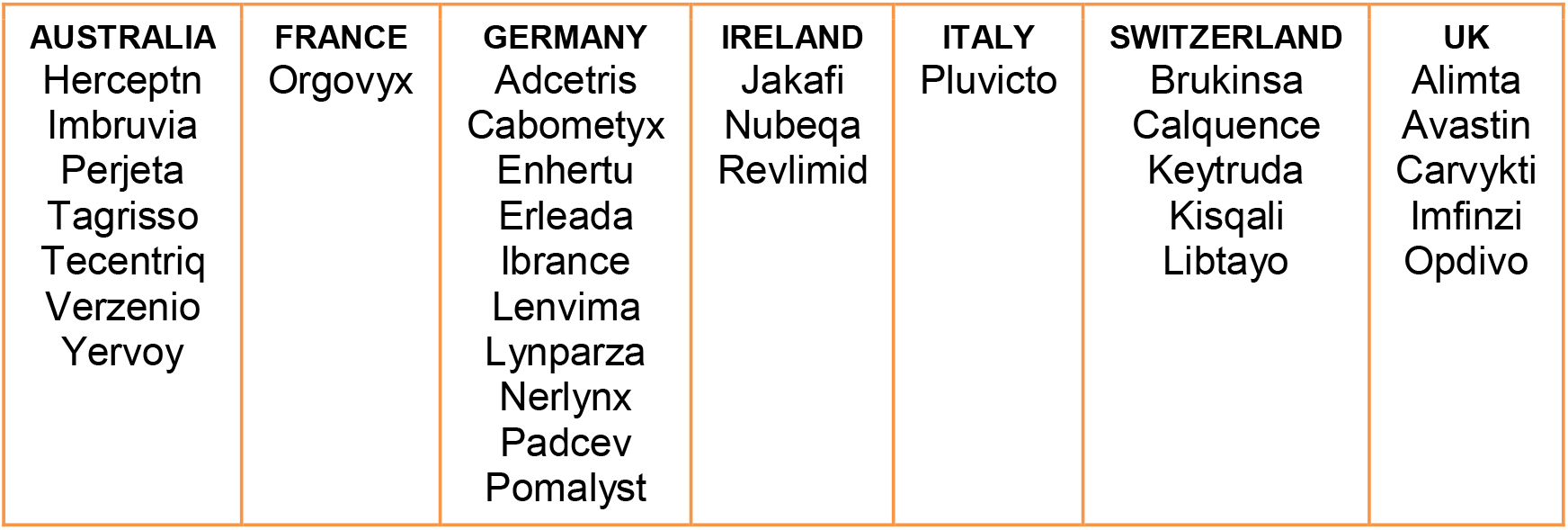
Cancer drug cohort, countries with the lowest GDP-adjusted price by drug.

Lead and secondary indications, dates of accelerated approvals, and dates of clinical trials were obtained from ClinicalTrials.gov and BioMedTracker, as well as secondary sources such as audited corporate reports and SEC filings.

Using calculated average costs per clinical trial participant from Joseph A. DiMasi et al.^8^, we conducted a retrospective analysis of these 37 FDA-approved cancer products to determine the impact of the theoretically lowest MFN price on the likelihood that each drug would have been developed had MFN been in place at the time of its FDA approval. Pre-clinical costs were applied based on their ratio to total net expenditure through phase III.^9^

Our revenue and cost data relate only to the lead indication; any revenue impact from post-approval indications was controlled through a regression analysis. Phase IV post-approval expenditures were excluded from our analysis. All monetary values were measured in constant 2024 USD; all cash flows were discounted using an 11% cost of capital.

To determine the impact of MFN on each drug, a NPV was modeled both for Medicare revenues in isolation and for Medicare prices ‘spilling over’ into the commercial market after a two-year delay (reflecting the likelihood that commercial payers in the US would demand MFN prices from pharmaceutical companies, leveraging payer coverage and formulary placement decisions).

An NPV is the value of all positive and negative cash flows over the entire life of an investment discounted to the present. NPV analysis is used extensively in biopharma investing, with the following standard formula:

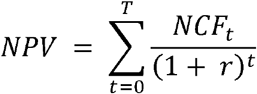

where NCF_t_ is the net cash flow at time t and r is the discount rate (i.e., cost of capital).

Lost future drugs, as determined by a negative NPV, were derived using the percent of cash sales allocated to R&D taken from research by NYU Stern Business School.^10^ The leading indications of all lost drugs were determined by US prevalence, and the number of potential patients losing access to medicines was calculated.

## Results

The average MFN net price reduction across our 37-drug cohort was 67%. Investors require a positive return on their investment. For 29 of our 37 approved cancer therapies, roughly 80%, had a positive NPV in the absence of MFN pricing (Figure 2) and were therefore ‘good’ investments.

**Figure 2:**
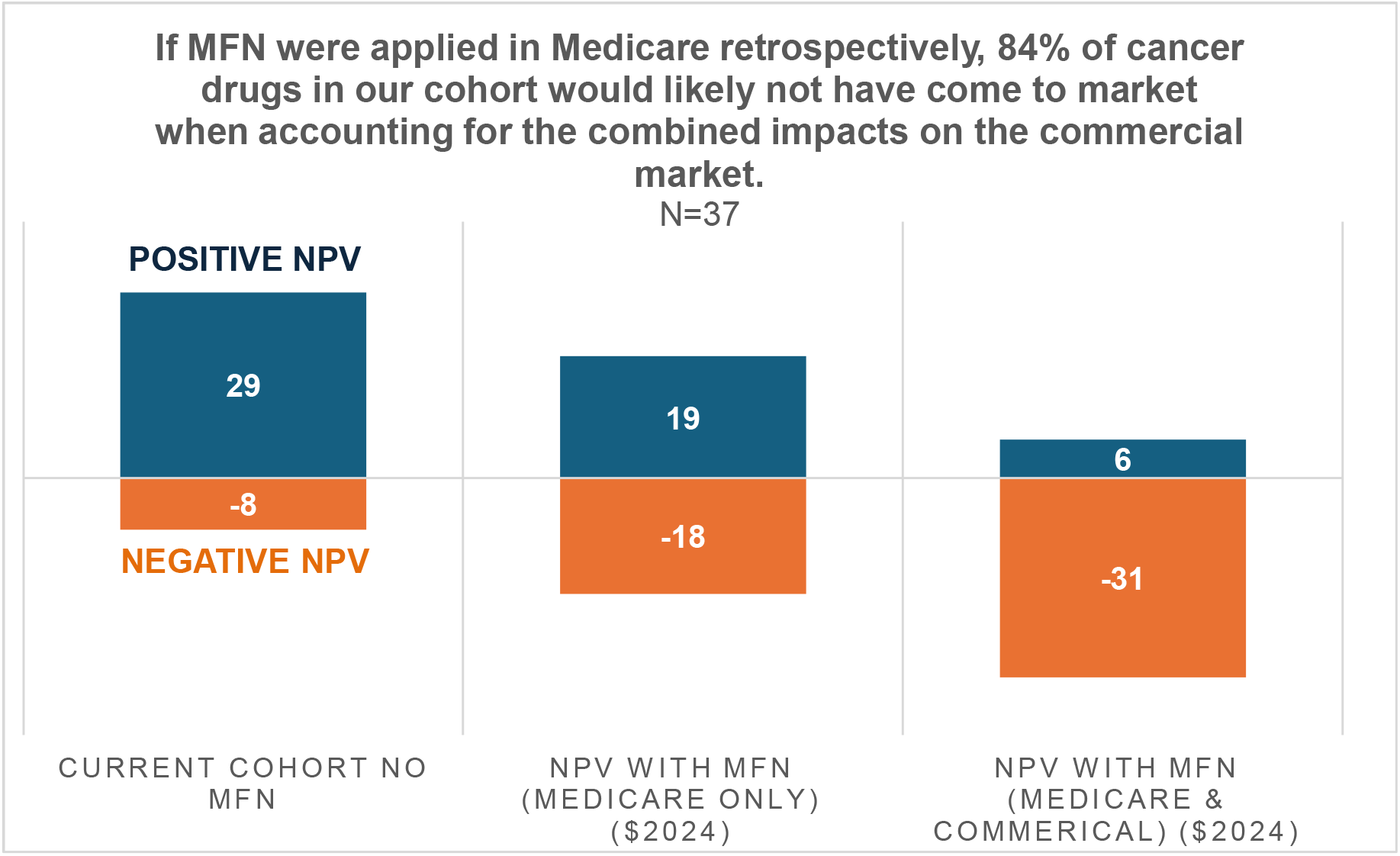
Impacts of MFN on the retrospective NPV of the current oncology cohort.

When MFN pricing is applied only to Medicare, roughly half (18 of 37) of previously approved drugs showed negative NPVs, meaning companies would likely not have supported their development had MFN been implemented at the time of approval.

As MFN price discounts can be extremely large, they are likely to also affect the commercial market. Assuming MFN prices affect the commercial market with a 2-year delay, we find that 84% (31 of 37) of the cancer drugs in our cohort negative NPVs and would therefore not likely have been developed had MFN been in place at the time of approval.

A cancer drug will be launched as a ‘lead’ indication and then be followed by testing on further types of cancer. If a drug’s lead indication cannot show a positive NPV, it will not be further tested for ‘secondary’ indications. For both small and large indications, MFN’s disincentives will impact patients’ access to needed new medicines (Figure 3).

**Figure 3:**
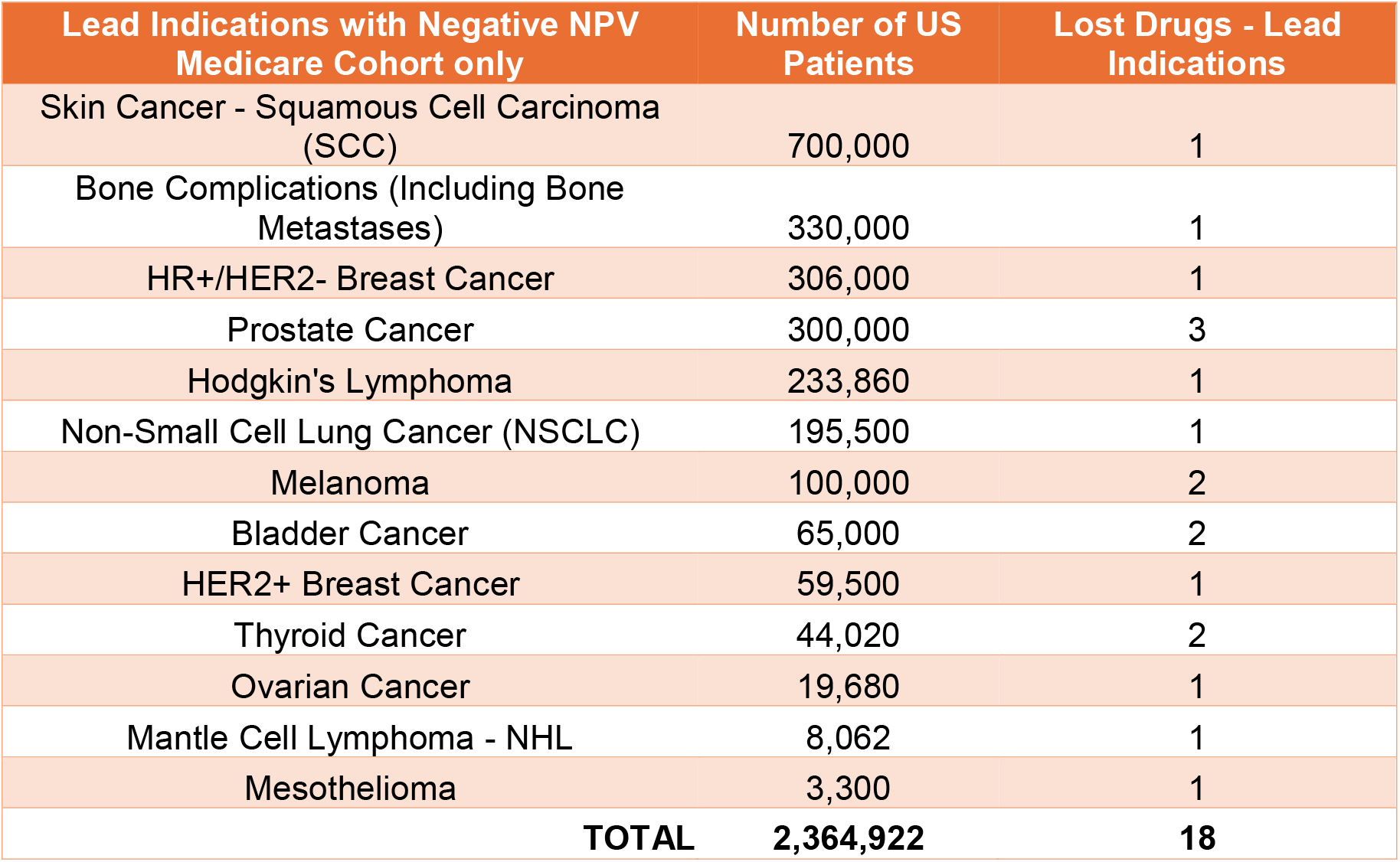
MFN’s impact restricted to Medicare, lead indications only. The total number of patients impacted represents the entire US cohort at risk if a new therapeutic is not brought to market; this may also be addressed by other therapies or by generic and biosimilar indications. It is indicative of the potential impact of losing a new, more effective drug.

If we restrict MFN’s impact to both Medicare and lead indications only, we find that the lead indications of the 18 therapies that no longer have a positive NPV address a total US population of 2.4 million cancer patients. Indications such as NSCLC often present late after metastasis has occurred, have a very high disease burden, and lack effective late-stage treatments.

When the impact of MFN also includes the commercial market, we see a far larger group of US patients potentially affected by the policy (Figure 4). We find that the lead indications of the 31 therapies that no longer have a positive NPV address a total US population of 15.2 million cancer patients, of which nearly 13 million receive a drug treating cancer-related anemia, which is no longer NPV-positive.

**Figure 4:**
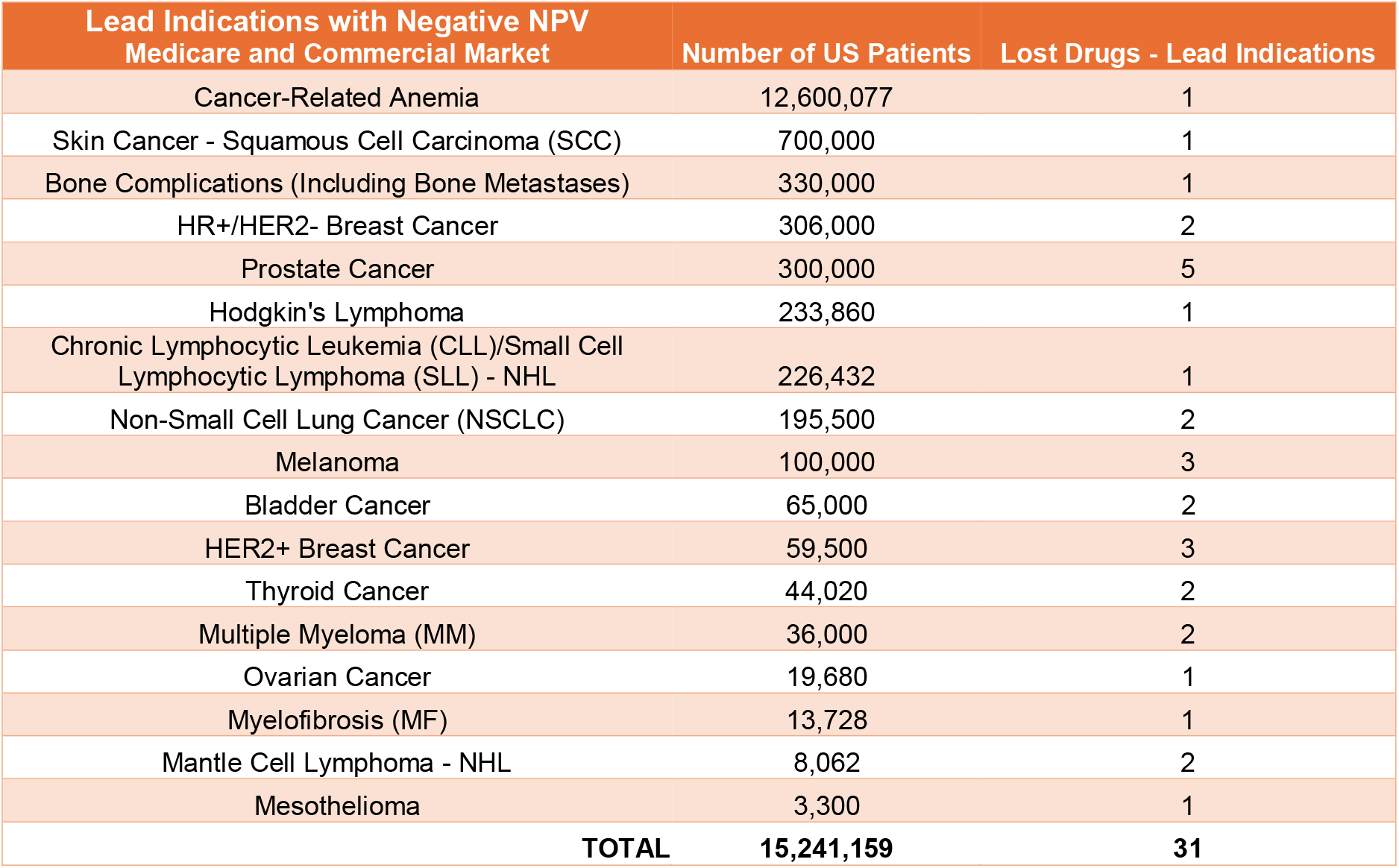
MFN’s impact in Medicare and commercial markets, lead indications only.

## Discussion

MFN pricing could have a demonstrable impact on US drug discovery and R&D, and it will be benchmarked to foreign prices set by mostly single-payer healthcare systems, where the government essentially acts as a monopoly buyer for all medicines. A recent study by IQVIA showed that the mean and median time to approval are 2.8 and 1.3 years faster, respectively, in the US compared to all EU 27 countries and Japan.^11^

The US provides greater access to more medicines. A 2018 study by the US Congress’ Ways and Means Committee found that, when a basket of 79 leading therapies available in the US was compared to their availability in similar OECD countries, the latter provided access to 25% fewer medicines.^12^ Further, U.S. payer decisions often support higher coverage and broader access than their foreign counterparts.^13^

Technically, MFN would peg US prices to the EU, which could lead to substantially lower sales volumes or an attempt to increase prices in Europe, despite that market’s sensitivity to higher prices. This could both impede patient access to new medicines in MFN countries and reduce the sector’s profitability. Lower revenues reduce available free cash flows for the development of new medicines.

The proposed GLOBE and GUARD demonstrations are involuntary, meaning that, if implemented, they would effectively be government-controlled prices at the MFN levels. Consensus in academic research concludes that the elasticity of pharmaceutical innovation is positive: as revenues fall (or rise), so does the degree of pharmaceutical innovation. Researchers at the USC Schaeffer Center conclude that for every 10% reduction in expected revenues, we can expect 2.5% to 15% less pharmaceutical innovation.^14^

Similarly, previous studies by the authors have shown that every 10% drop in the price of medicines in price-controlled EU markets is associated with a 14% decrease in total VC funding (10% early stage and 17% late stage), a 7% decrease in biotech patents, a 9% decrease in biotech start-up funding relative to the US, and an 8% increase in the delay of access to medicines.^15^ It therefore stands to reason that forcing similar price reductions in the US will have similar deleterious impacts, as seen in the EU, on the US biopharmaceutical ecosystem, with a direct beneficiary being Chinese innovation. Chinese companies currently conduct more clinical trials than companies based in the EU, and, on the current trend, they are set to eclipse those conducted by US companies by 2028.^16^

Finally, a prior study by the authors analyzing the response of venture capitalists to the Inflation Reduction Act^17^, which includes in its provisions other forms of government price controls on selected drugs^18^, found a statistically significant reduction in VC investments in small-molecule therapies targeting the Medicare-aged population; many being oncology therapies.^19^ MFN will only exacerbate this decline in the development and approval of new cancer medicines.

## Conclusion

If implemented, mandatory MFN policies could have a detrimental impact on patient access to pharmaceutical innovation in the US. Our examination of the potential impact on cancer drug development estimates a 50% to 84% decline in new cancer drug launches. When calculating MFN’s impact only in Medicare, the loss of lead indications could affect 2.4 million US patients. When MFN pricing is applied to both the Medicare and commercial markets, we find that the loss of lead and secondary indications could impact more than 15 million US patients.

The average reduction in US drug prices resulting from the MFN policy MFN was estimated to be 67%. Previous empirical studies have shown that price reductions have a statistically predictable impact on delays in access to medicines outside the US as well as on the number of drugs available.^20^

MFN reduces the financial incentives required to develop cancer medicines and will likely lead companies to pursue indications for populations outside Medicare’s authority.

If so, MFN will impact the very population the Executive Orders are allegedly designed to aid, namely, the Medicare-aged population who require effective new therapies in areas of high unmet medical need, such as late-stage cancers.

## Data Availability

Cohort data for this study is available at the following link:

https://1drv.ms/f/c/fd1ceff1664dae51/IgCoqrIcV_3FTo1zsjAqOvXaAWvX4hbvWRatfilNKRma6nc?e=45GEu5

## Funding

This research was funded by Eli Lilly. The authors received compensation as paid consultants to the sponsoring organization.

## Notes

### Competing Interest Statement

The authors received compensation as paid consultants to Eli Lilly, the sponsoring organization of this research.

